# Awareness, Acceptance and Associated Factors of Human Papillomavirus Vaccine Among Parents of Daughters in Hadiya Zone, Southern Ethiopia: A Cross-Sectional Study

**DOI:** 10.1101/2022.10.02.22280614

**Authors:** Yilma Markos Larebo, Legesse Tesfaye Elilo, Desta Erkalo Abame, Denebo Ersulo Akiso, Solomon Gebre Bawore, Abebe Alemu Anshebo, Natarajan Gopalan

## Abstract

**Introduction:** Human papillomavirus infections are the most prevalent sexually transmitted disease among women worldwide. Cervical cancer is the second most frequent disease worldwide in terms of incidence and mortality, and it is primarily responsible for fatalities in low- to middle-income nations, including Ethiopia. In the Hadiya Zone of Southern Ethiopia, the goal of this study is to determine parental knowledge about, acceptance of, and factors related to the human papillomavirus vaccine.

**Methods:** From November to December 2021, a community-based cross-sectional study would be conducted in Hadiya Zone among parents with daughters in the Zone. The study respondents would be chosen using a two-stage sampling technique from parents with a 9-14-year-old daughter. An interviewer-administered questionnaire would be used to collect data. For analysis, the data were entered into Epidata version 3.1 and exported to SPSS version 25. Variables with a p-value less than 0.25 in the bivariate analysis were transferred to multivariate analysis. A logistic regression model was applied to forecast the association between the predictor and outcome variables. Statistical significance was considered at a 0.05 p-value.

**Results:** The study showed that the overall acceptance of parents to vaccinate their daughter with HPV vaccination was 450 (84.9%). Parents of daughters having of male sex (AOR: 0.407; 95%CI: 0.221,0.748), having number of daughter’s one(AOR: 2.122; 95%CI: 1.221,3.685), government school type(AOR: 0.476;95%CI: 0.263,0.861), poor knowledge(AOR: 0.532;95%CI: 0.293,0.969) and negative attitude(AOR: 0.540;95%CI: 0.299,0.977) were discovered to have a strong correlation. **Conclusion:** This study found that there was a high level of parental acceptance; attitudes and knowledge about the HPV vaccine are significant in determining their intentions to vaccinate their daughter. Authorities in areas where cervical cancer incidence is at risk should plan and implement by providing health information regarding the human papillomavirus vaccination with an emphasis on raising community awareness.

## 1. Introduction

Human Papillomavirus (HPV) is the name given to a group of viruses that includes more than 200 related viruses, more than 40 of which are spread through vaginal and anal intercourse and can cause premalignant changes and malignant cancers of the cervix, vagina, vulva, and anus [1–4]. In both men and women, HPV infections are the most common sexually transmitted illness (STI)[1–3]. If an HPV infection goes untreated, it can lead to cancer and genital warts later in life [1,4–6].

Cervical cancer is the world’s most common cancer in women, every year, approximately 500 000 new cases [7–10], accounting for 7.5 %(270000) of leading causes of all female cancer death in women worldwide [8,11,12] and the malignant tumour arising from the cells of the uterine cervix[4]. It affects over a million women in the developed world, with HPV types 16 and 18 accounting for up to 70% of cases [5].

Cervical cancer results in a higher rate of cervical cancer death and the majority of deaths occur in low-to middle-income countries(LMICs) [4,7,12]. In LMICs with 70,000 new cases diagnosed each year more than 85 %(311 000) of the estimated deaths from cervical cancer, each year occur[11,12]. Sub-Saharan Africa (SSA) has the highest incidence and mortality rates in the world, accounting for more than 70% of the global cervical cancer burden[3,9].

In developing countries, the burden of 230,000 cases and deaths (80% of the total) is borne by, which have only the bare minimum of resources to deal with the situation[11,15,16], despite being largely preventable, it is the second most common cancer in developing countries in terms of incidence and mortality[4,7].

Cervical cancer is the second leading cause of cancer death among women in Ethiopia. Approximately 7600 new cervical cancer cases are diagnosed each year, with over 6000 deaths from the disease[7,17]. HPV is a major risk factor leading to the development of cervical cancer[1] and cervical cancer is caused by a persistent infection with one or more of the high-risk types of HPV[4,7,18],and it is the most common infection acquired during sexual relations, typically during childhood [11].

Cervical cancer screening is the most effective way for all women to reduce cancer mortality [7,12,17]. Although the majority of developed countries have well-organized screening, early detection, and effective treatment strategies for precancerous cervical lesions[3] more than 80% of sexually active women have HPV at some point in their lives[2].

As a result, the World Health Organization(WHO) develops comprehensive strategies for cervical cancer prevention and control, including HPV vaccination as the primary method of prevention[5]. Cervical cancer prevention programs, such as HPV vaccination and cytology-based cervical cancer screening programs have helped to reduce the incidence of the disease in developed countries[2].

Ethiopia introduced the HPV vaccine in 2018 with the help of the Global Alliance for Vaccine and Immunization (GAVI) [3,6,8].which was targeting 14-year-old girls and vaccines were administered through schools in two doses six months apart[17,20–22], but misconceptions about the cause and prevention of cervical cancer are common in Ethiopia due to a lack of awareness. As a result, appropriate studies to raise awareness, acceptance, and use of services such as immunization must be conducted before the nationwide scaling-up of cervical cancer prevention programs[2].

There is now an HPV prophylactic vaccine available that protects against most genital warts and cervical cancer. It is also known as the cervical cancer vaccine[1,5,17,23]. It accounted for approximately 90% of cervical cancer prevention cases and 10% of other HPV-related cancers or diseases[16,23,24]. HPV vaccines are generally regarded as safe, with only minor and transient side effects such as headache, dizziness, nausea, fatigue, soreness, redness, or swelling at the injection site[17,24–26].

The WHO recommends two doses of HPV vaccine for girls aged 9-14 years, and three doses for girls aged 15 years and older, as well as those who are immunocompromised or HIV positive (regardless of age)[12]. HPV vaccines work best for women who have never been exposed to HPV infection and are thus recommended for girls of appropriate ages before sexual debut; however, the vaccines do not prevent other sexually transmitted diseases or treat existing HPV infections or HPV-caused diseases[3,13]. It reduces/prevents infection with the following nine HPV types: HPV types 6 and 11, which cause 90% of genital warts; and HPV types 16 and 18, which cause approximately 70% of cervical cancers[4,18], but HPV types 31, 33, 45, 52, and 58, which cause 10% to 20% of cervical cancers[11,13,15].

Ethiopia had planned to introduce the HPV vaccine through a routine immunization program for approximately 6 million girls aged 9 to 14 years old[13,19]. However, due to a global shortage of HPV vaccine, the country is introducing the vaccine in a single age cohort (14-year-old girls) in the first year and hopes to expand the introduction to additional age cohorts in the second year and beyond based on global vaccine availability. If the vaccine shortage persists, the country will continue to vaccinate 14-year-old girls every year[19].

Basic knowledge of women’s pelvic anatomy and the natural history of cervical cancer provides primary and secondary level healthcare providers with the knowledge they need to effectively communicate and raise awareness of cervical cancer prevention in women, families, and communities[7]. Even if the study location is known to the investigators, there is little information about awareness, acceptance, and associated factors of the human papillomavirus vaccine among parents of daughters in the Hadiya Zone, Southern Ethiopia. Therefore, this study aimed to determine awareness, acceptance, and associated factors of the human papillomavirus vaccine among parents of daughters in the Hadiya Zone, Southern Ethiopia.

## 2. Methods and Materials

### 2.1. Study Setting, Design, and Period

A community-based cross-sectional study design was conducted in Hadiya Zone; among parents’ daughters of the Zone from November to December 2021. The Zone is located in South West of Ethiopia, 232 km far away from Addis Ababa(the capital city of Ethiopia), and 194 km from Hawassa (the regional capital city)[20,21].

Administratively, the Hadiya Zone was organized by 4 Administrative Town, 13-Districts, 305 rural Kebeles, and 30 urban Kebeles (smallest administrative unit of Ethiopia), with an estimated population size of 1,727,920 with male 856357(49.56%) and female 871563(50.44%)[20–22].

### 2.2. Sample Size Determination

The sample size was calculated using the single population proportion formula, considering the following assumptions and taking a prevalence of 79.5% which was a study conducted in the Bench-Sheko Zone, southwest Ethiopia[2], 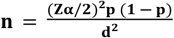Where n = the desired sample size P = parents’ acceptance to vaccinate their daughter= 79.5 % (which was taken from a study conducted in Bench-Sheko Zone, southwest Ethiopia,2021) Z1-α/2 = Critical value at 95% confidence level (1.96) d = the margin of error = 5% n = (<u>1.96)^2^ 0.795(1-0.795</u>) / (0.05)^2^ = 250 and using design effect (Deff=2), because two-stage sampling technique used, the final sample size required was: 2*250= 500, for possible none response during the study the final sample size, was increased by10% to: n =500+10% of 500 which is: 50, by adding then, the total sample size was 550.

### 2.3. Sampling Procedure

The two-stage sampling technique was used to select study respondents. At the first stage of sampling, Kebeles would be stratified as urban or rural, and two urban and ten rural Kebeles would be chosen using a simple random sampling technique. In the second stage, four Districts Lemo, Misha, Soro, and Shashogo as well as the Hossana and Shone town administrations were chosen by lottery methods. Then, thirty Percent of Kebeles from each of the selected Districts were chosen.

The calculated samples would be then distributed to each of the selected schools in Kebeles under the proportional to sample size allocation (PPS) principle. Respondents would be parents or caregivers or guardians of a daughter aged 9 to 14, such as parents, grandparents, or anyone else who has self-identified as being responsible for the daughter’s care. Male caregivers would be interviewed if a female caregiver is unavailable. Female caregivers would be preferred. If a household has more than one female between the ages of 9 and 14, one would be chosen at random as the index female for interview questions. Finally, the 550 daughters would be chosen using a systematic random sampling technique with a skip interval of every ‘5th’. As a result, households with daughters aged 9–14 would be interviewed.

Parents or guardians of daughters aged ≥ 18 years old who live permanently in the study area (for more than 6 months) were included in the study. Those respondents who were unable to provide the necessary information, pregnant girls, girls who had recently given birth and were breastfeeding, and girls who were not attending classes despite being enrolled, were also excluded, and could not be recruited. When no person could participate in the study, the next household was used in their place.

### 2.4. Measurement of Variables

The purpose of this study was to determine awareness, acceptance, and associated factors of the human papillomavirus vaccine among parents of daughters. The dependent variable in this study was acceptance of the human papillomavirus vaccine. The predictor variables were respondents’ basic socio-demographic characteristics such as age, sex, educational status, marital status, occupational status, monthly income level, knowledge, attitude, and awareness of HPV vaccination.

In this study, the knowledge of the parents of daughters on acceptance of the human papillomavirus vaccine should be measured like 17 yes or no response items questioner would be used. A correct response was assigned a value of one, while an incorrect response was assigned a value of zero. The scores for each item are added up, and those who scored higher than the mean value are classified as having good knowledge, while those who scored lower are classified as having poor knowledge[2,4,18].

The Attitude of the parents of daughters on acceptance of the human papillomavirus vaccine should be determined by eleven Likert scale items ranging from strongly disagree to strongly agree will be used to assess attitude. Those who scored higher than the mean value were classified as having a positive attitude, while those who scored lower were classified as having a negative attitude[2,4,18].

The acceptance of the parents of their daughter**’s** acceptance of the human papillomavirus vaccine all respondents, the question “are you willing to vaccinate your daughter for HPV vaccination, which can protect against HPV infection?” would be used to assess acceptance (Choices: 1 as Yes or 0 as No)[2,4,18].

The awareness of the parents of daughters on acceptance of the human papillomavirus vaccine should be measured like the question today, had you heard of HPV? be applied to assess the awareness of HPV. Parents of daughters who answered ‘yes’ to this question would be regarded to be aware of HPV[2,4,18].

### 2.5. Data Collection and Quality Assurance Procedures

The data were collected using interviewer-administered quantitative data. The questionnaire contains questions developed and adapted from various kinds of literature. The necessary permissions were obtained from the owner of the original questionnaire. The questionnaire developed by the investigators contained the following 4 sections: 1 basic demographic characteristic (age, sex, educational status, marital status, occupational status, and monthly income level), 2 knowledge, 3 attitude, and 4 information-related factors on vaccination awareness and acceptance of parents of daughters on acceptance of human papillomavirus vaccine. Three days of training were provided for data collected using an interview-administered method by two trained data collectors and one supervisor. The principal investigator reviewed the obtained data to ensure its accuracy, completeness, clarity, and consistency. The questionnaire was written in English, translated into common Amharic to ensure that the items were consistent, and then translated back into English to ensure that the common Amharic was accurate. The questionnaire’s face validity was established.

A pre-test was performed before data collection by taking 28 samples out of the study area. The data collection tool’s reliability would be evaluated in terms of knowledge, attitude, acceptance as well as awareness. To ensure the quality of the data, emphasis was placed on the design of the data collection instrument for its simplicity and standardized community rating scales, validity and reliability would be considered.

### 2.6. Data Processing and Analysis

Data that had been coded was input in Epi Data version 3.1 and exported to SPSS version 25 for analysis. Data entry fell under the purview of the principal investigator. Tables, graphs, and charts were used to do the descriptive analysis and report the results in frequency.

Variables from the bivariate analysis that had P-values under 0.25 were added to the multivariate analysis.

To choose variables for multivariate analysis, bivariate analysis was used. However, multivariate statistical significance was tested at the level of 5%. Using logistic regression, adjusted odds ratios (AOR) and a 95% confidence interval were utilized to confirm the presence and strength of the link between the independent and dependent variables. The 0.816 Hosmer and Lemeshow tests were used to gauge the model’s fitness. The Variance Inflator Factor (VIF), standard error, and correlation coefficient would be used to test for multicollinearity among independently associated variables.

### 2.7. Ethical Approval

The WCU College of Medicine and Health Sciences ethical review committee (ERC) provided the necessary ethical approval. Permission from the District educational offices, schools, and Zone-education Department was secured before the study began. Before being enrolled, respondents received a written agreement that included information about the study’s aims, consequences, and the importance of the data. All information was made anonymous to maintain confidentiality

## 3. Results

### 3.1. Characteristics of Respondents

Out of the 550 parents of daughters who were eligible to participate in the study, 20 were excluded (20 data were incomplete and were not considered for analysis), resulting in a 96.54% response rate. Out of 530 parents of daughters included in the study, the majority 323(60.9%) of the respondents were female, and almost more than half 323(60.9%) of them were in the age category 30-39 years. The mean age of parents of daughters was 38 with (SD) + (9.45) years and nearly less than three fourth 365(68.9%) of the respondents were protestant, 354(66.8%) were urban by residence, 437(82.5%) were married by marital status, 322(60.8%) were Degree and above by education status, 354(66.8%) were Civil servant by occupation and 389(73.4%) were Hadiya ethnicity. The majority of the respondents 418(79.8%) had more than one child, 355(67%) respondents were learn in a private school, more than half of respondents318(60%) have information on HPV and 144(45.3%) were the major source of information Television or Radio. Almost half 275(51.9%) respondents were monthly income >4999 ETB (≤95.24USD) (Table 1).

**Table 2.** Sociodemographic characteristics of the respondents on awareness, acceptance, and associated factors of human papillomavirus vaccine among parents of daughters in Hadiya Zone, Southern Ethiopia, 2021 (n=530).

| Variables | Categories | n (%) |
| --- | --- | --- |
| Sex | Male | 207(39.1) |
|  | Female | 323(60.9) |
| Age in years | 18-29 | 80(15.1) |
|  | 30 to 39 | 323(60.9) |
| | ≥40 | 127(24) |
| Residence | Rural | 176(33.2) |
|  | Urban | 354(66.8) |
| What is your marital status now | Single | 69(13) |
|  | Married | 437(82.5) |
|  | Ever married | 24(4.5) |
| Religion | Orthodox | 86(16.2) |
|  | Muslim | 64(12.1) |
|  | Protestant | 365(68.9) |
|  | catholic | 15(2.8) |
| Educational status | Unable to read and write | 16(3) |
|  | Primary and secondary education | 96(18.1) |
|  | certificate | 32(6) |
|  | Diploma | 64(12.1) |
|  | Degree and above | 322(60.8) |
| Occupation | Civil servant | 354(66.8) |
|  | Self-employers | 80(15.1) |
|  | Merchant | 80(15.1) |
|  | Housewife | 16(3) |
| Ethnicity | Hadiya | 389(73.4) |
|  | Kembata | 50(9.4) |
|  | Amhara | 8(1.5) |
|  | Tigre | 16(2.1) |
|  | Gurage | 10(1.9) |
|  | Oromo | 9(1.7) |
|  | Wolaita | 16(3) |
|  | Gurage | 32(6) |
|  | Silte | 16(3) |
|  | others* | 41(5.4) |
| Number of daughters | One | 112(21.1) |
|  | More than one | 418(79.8) |
| School type | Government School | 175(33) |
|  | Private school | 355(67) |
| Information about HPV infection | Yes | 318(60) |
|  | No | 212(40) |
| Sources of information about HPV infection(more than one possible answer possible) | Television(TV) or Radio | 144(45.3) |
|  | Health workers | 78(24.5) |
|  | Their friend | 70(22) |
|  | Others** | 26(8.2) |
| Monthly income | ≤4999 ETB (≤95.24USD) | 239(45.1) |
|  | >4999 ETB (>95.24 USD) | 291(54.9) |

Where: TV; Television, ETB; Ethiopian Total Birr, USD; United States Dollars \*=23 Alaba,
6 Sidama, 12 Dawro, \*\*=12 internets, 5 email messages, 7 groceries, and 2 newspapers.

Of the 530 study respondents, the majority, 419(79.1%) with 95% CI [75.5, 82.5] had good knowledge with a mean of 1.7906 and standard deviation of ±0.40729 (Fig 1)

**Figure 1.**
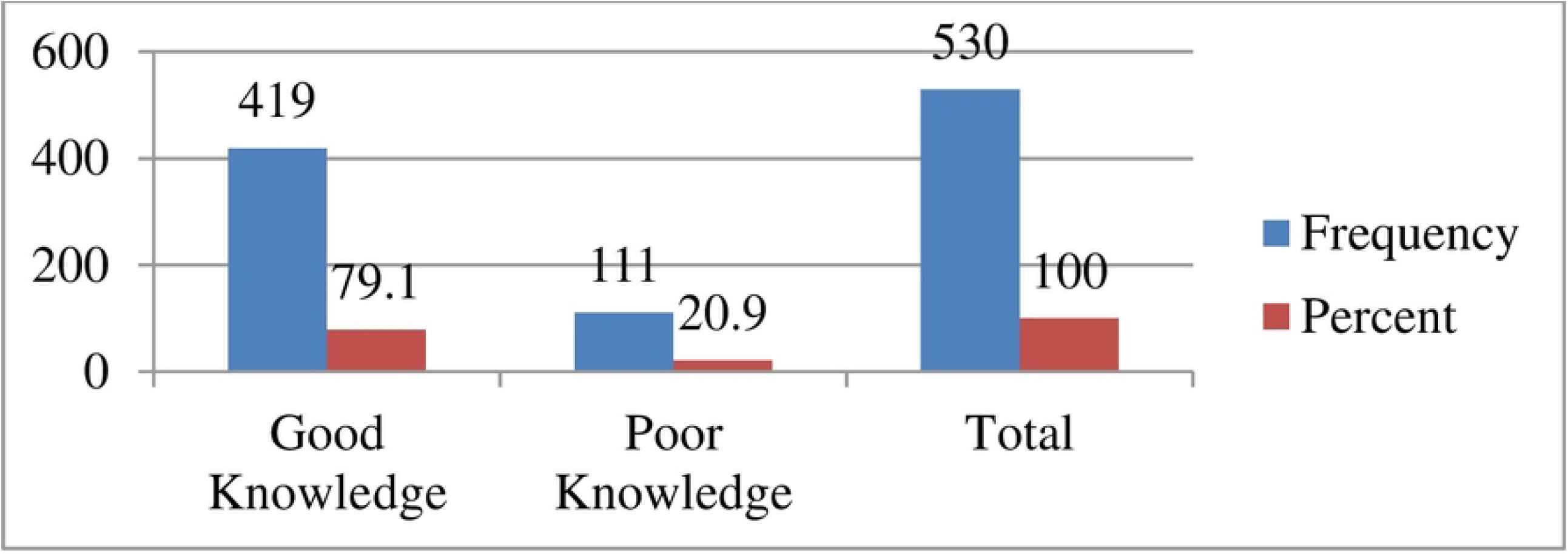
Knowledge-related questionnaire of the respondents on awareness, acceptance, and associated factors of human papillomavirus vaccine among parents of daughters in Hadiya Zone, Southern Ethiopia, 2021 (n=530).

Almost more than three fourth 418(78.9%) and nearly all most all 499(94.2%) of the parents of daughters ever heard about cervical cancer and cervical cancer infect only females, respectively. Nearly less than one-fourth, 111(20.9%) and 144(27.2%) of the parents of daughters know about cervical cancer at an early stage produces no signs and symptoms and it is fast-growing cancer, respectively. Concerning the treatment curability, about 418(78.9%) respondents’ answers the correct answers by early detection for cervical cancer protection and 371(70%) respondents were getting a Pap test for early detection of cervical cancer. Almost three fourth 387(73%) respondents knew about HPV is the main cause of cervical cancer and were majority 435(82.1%) common in women younger than 30 years old. More than three fourth 419(79.1%) and 434(81.9%) respondents knew about HPV vaccination is available for girls aged 9 to 14 years and cervical cancer risk can be reduced by HPV vaccination, respectively. Almost more than half of 307(57.9%) and 338(63.8%) of the respondents knew that HPV infection is a sexually transmitted infection and vaccination should be received before sexual debut, respectively. The majority 434(81.9%) and nearly less than one-fourth of 112(21.1%) of the respondents knew that the persistent infection of high-risk HPV could lead to cervical cancer and 70% of cervical cancer is caused by HPV types 16 and 18, respectively. Most more than one-fourth 159(30%) of the parents of daughters were they know the common and effective treatment to cure cervical cancer (Table 2).

**Table 2.** Knowledge-related questionnaire of the respondents on awareness, acceptance, and associated factors of human papillomavirus vaccine among parents of daughters in Hadiya zone, southern Ethiopia, 2021 (n=530).

| Variables | Categories | n (%) |
| --- | --- | --- |
| Have you ever heard about cervical cancer? | Yes | 418(78.9) |
|  | No | 112(21.1) |
| Does cervical cancer infect only females? | Yes | 499(94.2) |
|  | No | 31(5.8) |
| Cervical cancer at an early stage produces no signs or symptoms. | yes | 111(20.9) |
|  | No | 419(79.1) |
| Cervical cancer is fast-growing cancer. | Yes | 144(27.2) |
|  | No | 386(72.8) |
| If detected early, is cervical cancer curable? | Yes | 418(78.9) |
|  | No | 112(21.9) |
| Getting a Pap test helps early detection of cervical cancer | Yes | 371(70) |
|  | No | 159(30) |
| Human papillomavirus is the main cause of cervical cancer | Yes | 387(73) |
|  | No | 143(27) |
| Human papillomavirus is very common in women younger than 30 years. | Yes | 435(82.1) |
|  | No | 95(17.9) |
| Human papillomavirus vaccination is available for girls aged 9 to 14 years. | Yes | 419(79.1) |
|  | No | 111(20.9) |
| Cervical cancer risk can be reduced by HPV vaccination. | Yes | 434(81.9) |
|  | No | 96(18.1) |
| Do you know that HPV vaccination should be received before sexual debut | Yes | 338(63.8) |
|  | No | 192(36.7) |
| Know that HPV infection is a sexually transmitted infection | Yes | 307(57.9) |
|  | No | 223(42.1) |
| Know that the persistent infection of high-risk HPV could lead to cervical cancer | Yes | 434(81.9) |
|  | No | 96(18.1) |
| Do you know the symptoms of cervical cancer? | Yes | 386(72.8) |
|  | No | 144(27.2) |
| Do you know the time duration that takes for the abnormal cervical cells to turn into cancerous cells? | Yes | 80(15.1) |
|  | No | 450(84.9) |
| 70% of cervical cancer is caused by HPV types 16 and 18. | Yes | 112(21.1) |
|  | No | 418(78.9) |
| Do you know the common and effective treatment to cure cervical cancer? | Yes | 159(30) |
|  | No | 371(70) |

Where: HPV, Human papillomavirus.

Out of 530 total respondents, 371 (7 0%) had positive attitudes with a 95 % CI [65.9, 74] and a mean of 1.7000, and a standard deviation of ± 0.45869(Fig 2).

**Figure 2.**
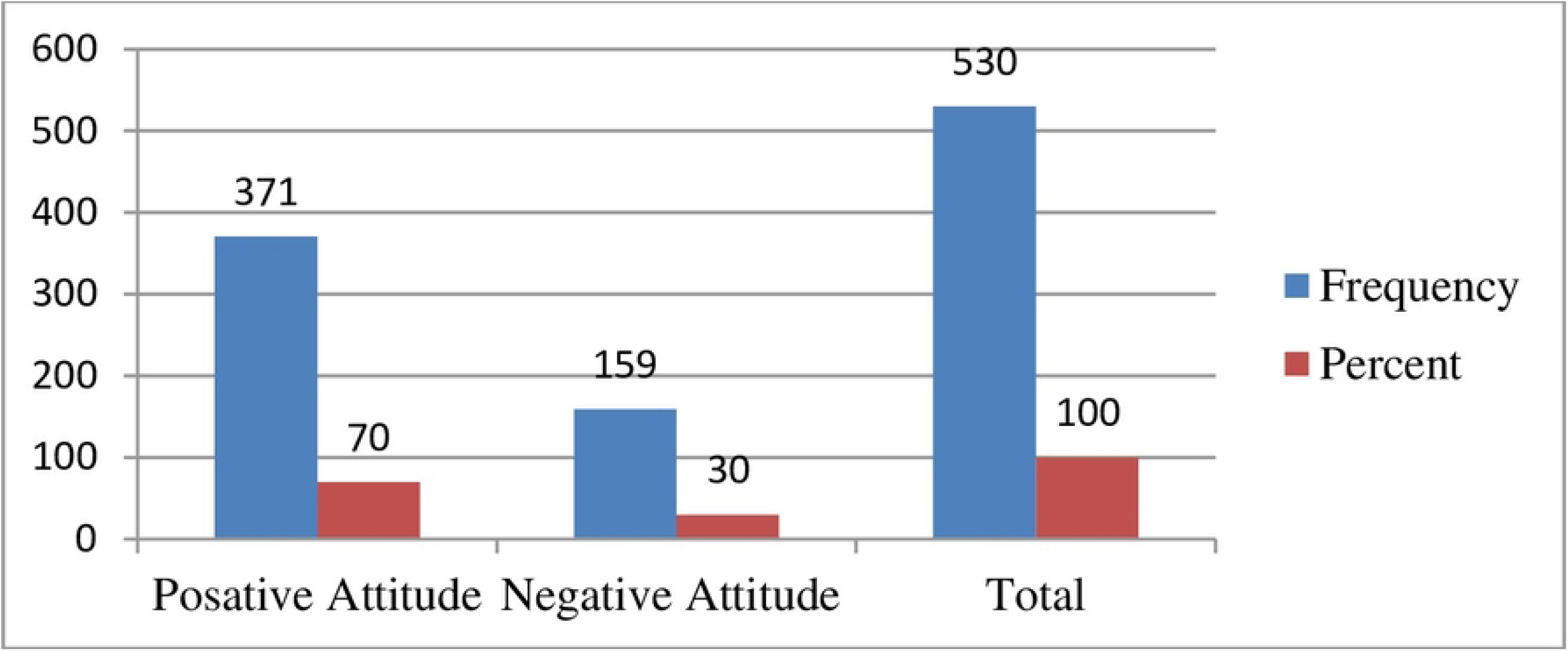
Attitude-related questionnaire of the respondents on awareness, acceptance, and associated factors of human papillomavirus vaccine among parents of daughters in Hadiya zone, southern Ethiopia, 2021 (n=530).

This study identified more than half of 291 (54.9%) and nearly three fourth 387(73%) of the daughters of parents were willing to regularly consult a medical doctor for cervical cancer screening and those daughters with multiple sex partners will be at higher risk for cervical cancer, respectively. The majority 413 (77.9%) and 386 (72.8%) of parents of daughters agreed that long-term use of contraceptive pills could cause and also the use of condoms could reduce the risk of cervical cancer respectively. More than half 370 (69.8%) of the respondents were doesn’t consult a medical doctor in case of abnormal bleeding between menstrual periods while 386 (72.8%) of parents of daughters agreed that HPV vaccination can prevent cervical cancers. The majority 434(81.9%) and 483(91%) were willing to get a Pap smear test and a regular Pap smear test helps early detection of cervical cancer respectively. The majority of 467(88.1) respondents disagreed to pay for a Pap smear test and 463 (87.4%) respondents agreed that the government should provide free screening programs to reduce cervical cancer prevalence. The majority 434(81.9%) parents of daughters have disagreed that the HPV vaccine might cause short-term problems, like fever or discomfort (Table 3).

**Table 3.** Attitude-related questionnaire of the respondents on awareness, acceptance, and associated factors of human papillomavirus vaccine among parents of daughters in Hadiya zone, southern Ethiopia, 2021(n=530).

|  | n | % | n | % | n | % | n | % | n | % |
| --- | --- | --- | --- | --- | --- | --- | --- | --- | --- | --- |
| Are you willing to regularly consult a medical doctor for cervical cancer screening? | 175 | 33 | 16 | 3 | 48 | 9.1 | 291 | 54.9 | 0 | 0 |
| Those with multiple sex partners will be at higher risk for cervical cancer | 64 | 12.1 | 15 | 2.8 | 64 | 12.1 | 371 | 70 | 16 | 3 |
| Long-term use of contraceptive pills could cause cervical cancer | 112 | 21.1 | 29<br>6 | 55.8 | 5 | 0.9 | 90 | 17 | 27 | 5.1 |
| The use of condoms could reduce the risk of cervical cancer | 16 | 3 | 15 | 2.8 | 80 | 15.1 | 64 | 12.1 | 355 | 67 |
| HPV vaccination can prevent cervical cancers | 48 | 9.1 | 32 | 6 | 64 | 12.1 | 80 | 15.1 | 306 | 57.7 |
| Will you consult a medical doctor in case of abnormal bleeding between menstrual periods? | 274 | 51.7 | 16 | 3 | 80 | 15.1 | 80 | 15.1 | 80 | 15.1 |
| Regular Pap smear test helps early detection of cervical cancer | 16 | 3 | 16 | 3 | 15 | 2.8 | 355 | 67 | 128 | 24.2 |
| Are you willing to get a Pap smear test? | 32 | 6 | 32 | 6 | 32 | 6 | 111 | 20.9 | 323 | 60.9 |
| Are you willing to pay for a Pap smear test? | 64 | 12.1 | 64 | 12.1 | 33<br>9 | 64 | 32 | 6 | 31 | 5.8 |
| Government should provide free screening programs to reduce cervical cancer prevalence. | 32 | 6 | 31 | 5.8 | 7 | 1.3 | 364 | 68.7 | 96 | 18.1 |
| The HPV vaccine might cause short-term problems, like fever or discomfort | 16 | 3 | 32 | 6 | 38<br>6 | 72.8 | 48 | 9.1 | 48 | 9.1 |

Out of 530 total respondents, 450 (84.9%) parents were accepted to vaccinate their daughter for HPV vaccination with a 95 % CI [81.9, 87.9] and a mean of 1.1509 and a standard deviation of ± 0.35833 (Fig3) and obstacles(Fig4).

**Figure 3.**
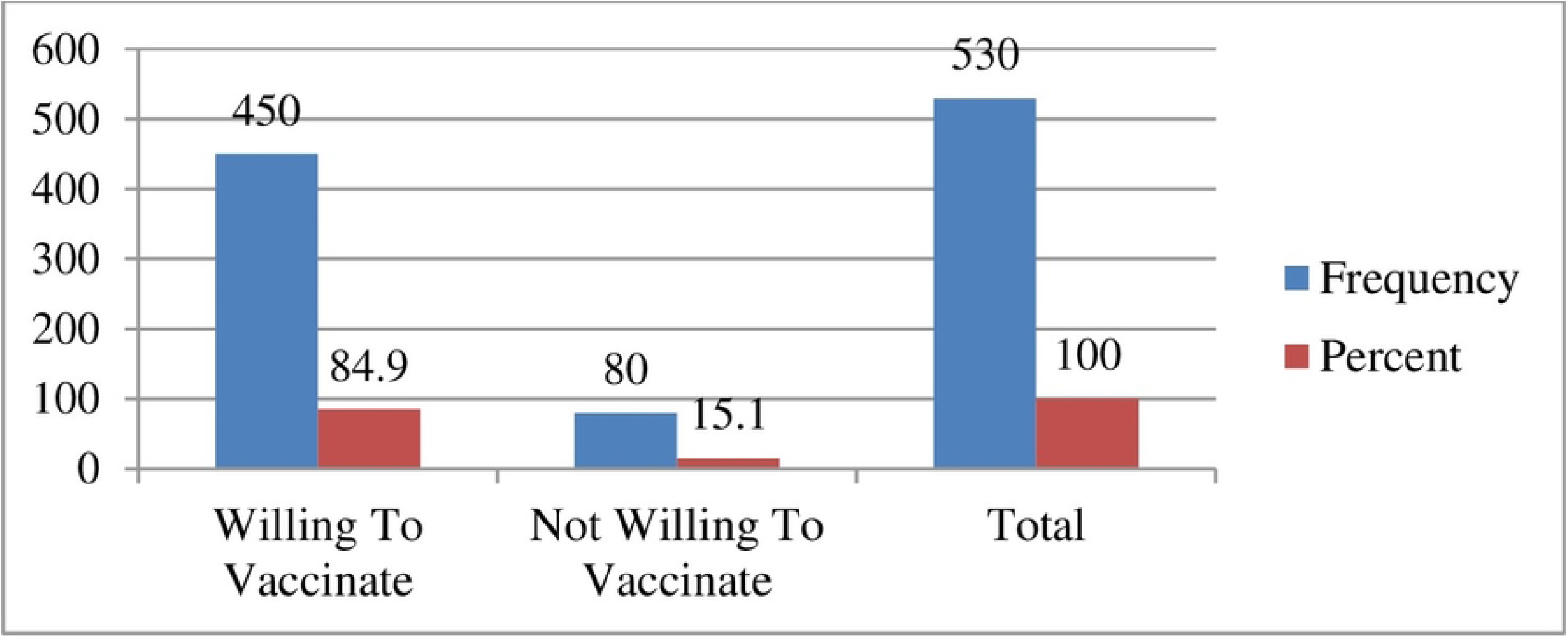
Acceptance-related questionnaire of the respondents on awareness, acceptance, and associated factors of human papillomavirus vaccine among parents of daughters in Hadiya zone, southern Ethiopia, 2021 (n=530).

**Figure 4.**
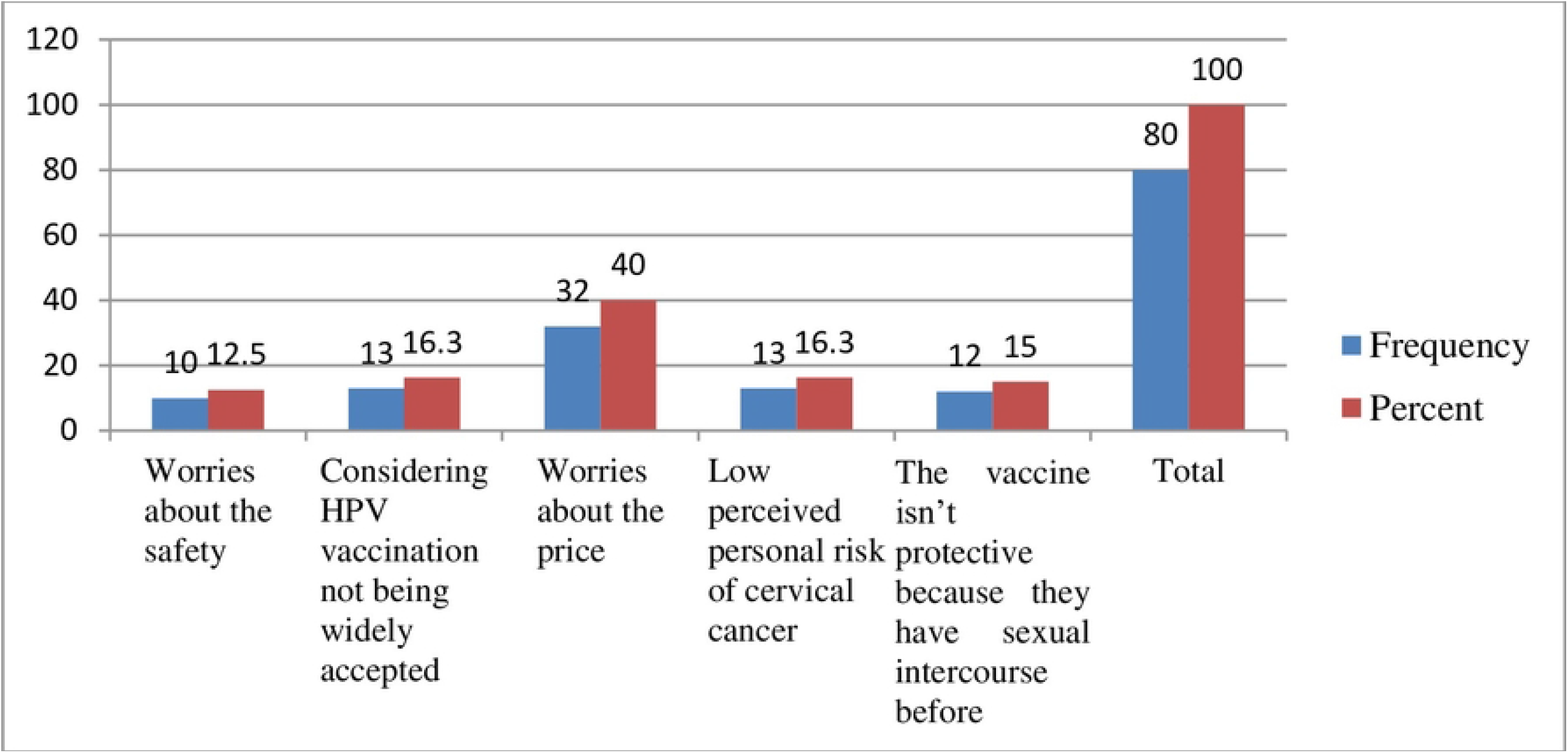
Obstacles to the acceptance-related questionnaire of the respondents on awareness, acceptance, and associated factors of human papillomavirus vaccine among parents of daughters in Hadiya zone, southern Ethiopia, 2021 (n=530).

Out of 530 total respondents, 450 (84.9%) parents were accepted to vaccinate their daughter for HPV vaccination with a 95 % CI [81.7, 88.1] and a mean of 1.1509 and a standard deviation of ± 0.35833 (Fig 5).

**Figure 5.**
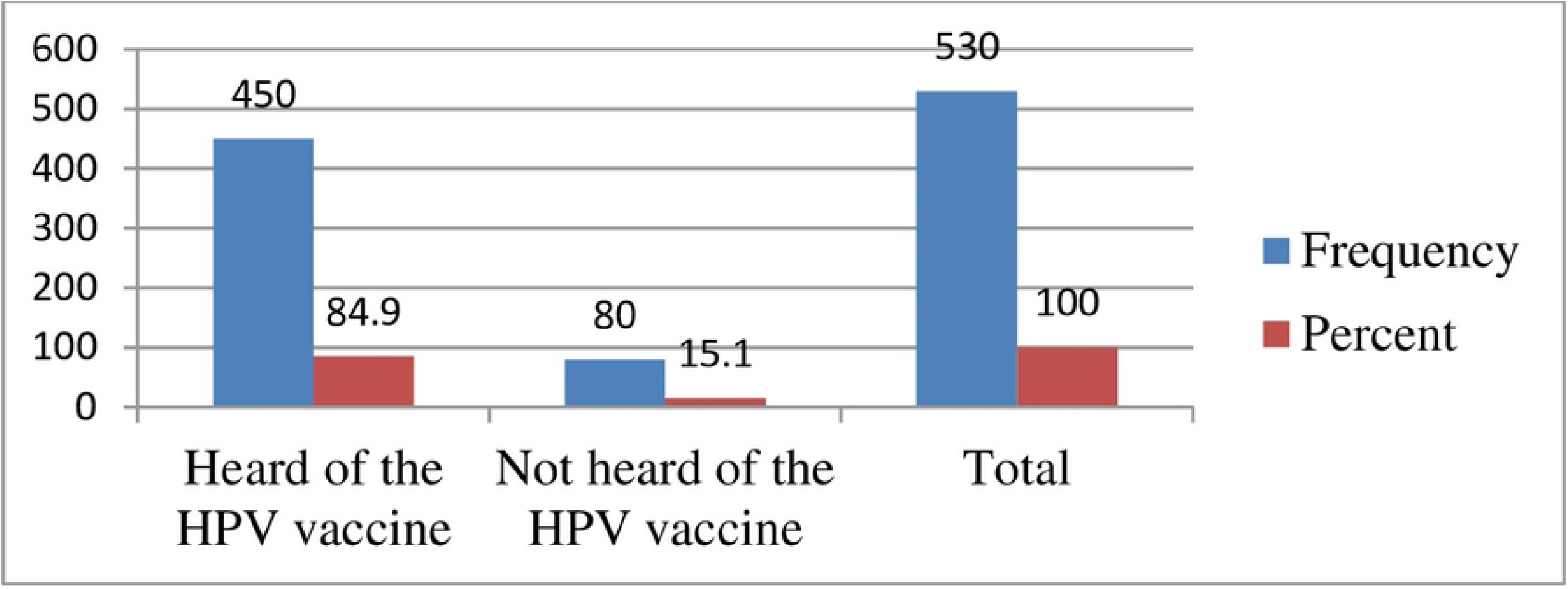
Awareness-related questionnaire of the respondents on awareness, acceptance, and associated factors of human papillomavirus vaccine among parents of daughters in Hadiya zone, southern Ethiopia, 2021 (n=530).

The majority 467(88.1%) and 450(84.9%) of the parents of daughters heard and vaccinate about HPV respectively. The majority 450(84.9%) of the study respondents were willing to accept to vaccinate their daughter with HPV vaccination and 32(6%) reasons for being unwilling to take the HPV vaccine worried about the price and the majority 482(90.9%) respondents were they do not accept that they pay for the HPV vaccination by themselves(Table 4).

**Table 4.** Acceptance and awareness -related questionnaire of the respondents on awareness, acceptance, and associated factors of human papillomavirus vaccine among parents of daughters in Hadiya zone, southern Ethiopia, 2021 (n=530).

| Variables | Categories | n (%) |
| --- | --- | --- |
| Heard of HPV | Yes | 467(88.1) |
|  | No | 63(11.9) |
| Heard of the HPV vaccine | Yes | 450(84.9) |
|  | No | 80(15.1) |
| Are you willing to vaccinate your daughter for HPV vaccination? | Yes | 450(84.9) |
|  | No | 80(15.1) |
| The reasons for being unwilling to take the HPV vaccine (more than one possible answer possible (obstacles)). | Worries about the safety | 10(1.9) |
|  | Considering HPV vaccination not being widely accepted | 13(2.5) |
|  | Worries about the price | 32(6) |
|  | Low perceived personal risk of cervical cancer | 13(2.5) |
|  | The vaccine isn't protective because they have sexual intercourse before | 12(2.3) |
| Accept that they pay for the HPV vaccination by themselves | Yes | 48(9.1) |
|  | No | 482(90.9) |

#### Factors Associated With the Acceptance of Human Papillomavirus Vaccine among Parents of Daughters

Multivariable analysis was used to control potential confounders. Having parents of daughters of the male sex, having a number of daughters, government school type, poor knowledge, and having a negative attitude were discovered to have a substantial relationship with acceptance of the human papillomavirus vaccine with a P-value < 0.05.

In this study, parents of daughters of the male sex were 59.3% less likely to accept HPV vaccination for their daughters as compared to the female sex (AOR: 0.407; 95%CI: 0.221,0.748), having a number of daughters one were 2.122 times more likely to accept HPV vaccination for their daughters as compared to the number of daughters more than one (AOR: 2.122; 95%CI: 1.221,3.685), parents of daughters having government school type were 52.4% less likely to accept HPV vaccination for their daughters as compared to private school (AOR: 0.476;95%CI: 0.263,0.861), parents of daughters having poor knowledge were 46.8% less likely to accept HPV vaccination for their daughters as compared to good knowledge(AOR: 0.532;95%CI: 0.293,0.969) and those parents of daughters having negative attitude were 46% times less likely to accept HPV vaccination for their daughters as compared to positive attitudes (AOR: 0.540;95%CI: 0.299,0.977) (Table 5).

**Table 5.** Multivariate And bivariate Analyses of Factors Associated with the Acceptance of Human Papillomavirus Vaccine among Parents of Daughters Hadiya Zone, Southern Ethiopia, 2021 (n=530).

| Variable | Acceptability of the HPV Vaccination |  |  |  |  |
| --- | --- | --- | --- | --- | --- |
|  | No | Yes | COR(95%CI) | AOR(95%CI) | P-value |
| Sex |  |  |  |  |  |
| Male | 48(60) | 159(35.3) | 0.364(0.224,0.539) | 0.407(0.221,0.748)* | 0.004 |
| Female | 32(40) | 291(64.7) | 1 | 1 |  |
| Number of daughters |  |  |  |  |  |
| One | 32(40) | 80(17.8) | 0.324(0.195,0.539) | 2.122(1.221,3.685)* | 0.008 |
| More than one | 48(60) | 370(82.2) | 1 | 1 |  |
| School type |  |  |  |  |  |
| Government School | 48(60) | 127(28.2) | 0.262(0.160,0.429) | 0.476(0.263,0.861)* | 0.014 |
| Private school | 32(40) | 323(71.8) | 1 | 1 |  |
| Knowledge |  |  |  |  |  |
| Poor knowledge | 32(40) | 79(17.6) | 0.319(0.192,0.531) | 0.532(0.293,0.969)* | 0.039 |
| Good knowledge | 48(60) | 371(82.4) | 1 | 1 |  |
| Attitude |  |  |  |  |  |
| Negative Attitude | 32(40) | 127(28.2) | 0.590(0.361,0.965) | 0.540(0.299,0.977)* | 0.042 |
| Positive Attitude | 48(60) | 323(71.8) | 1 | 1 |  |

Where 1 =Reference,\* shows the variable significant at p-value < 0.05 in multi variable analysis.

## 4. Discussion

This study was conducted on parents of daughters attending school to see if a structured educational intervention could improve parental awareness, knowledge, attitudes, and views regarding HPV infection, cervical cancer, and HPV vaccination as well as the acceptance of the HPV vaccine. The major purpose of this study was to determine parental awareness, acceptance, and associated factors of the human papillomavirus vaccine in the Hadiya zone, southern Ethiopia. HPV prophylactic vaccine is available that protects against most genital warts and cervical cancer and is known as the cervical cancer vaccine[1,12,16,23]. It accounted for approximately 90% of cervical cancer prevention cases and 10% of other HPV-related cancers or diseases[15,16].

The main finding of this study revealed that, despite respondents’ restricted tolerance for male sex and government-school knowledge about the HPV vaccine, the majority of them were willing to immunize their children. This study finding is also almost nearly two times more than the study conducted in Hong Kong[23], and 1.12 times more than in Lagos, Nigeria[24].

According to the findings, the overall acceptance of HPV vaccinations for their daughters was 450 (84.9%), when compared to a previous study conducted 3.6 times higher than the study conducted among male baccalaureate students in Hong Kong[1], 1.1 times higher in Gondar town, Northwest Ethiopia[18], 2.12 times higher when the study conducted in the United States[16], 1.24 times more than the study conducted a premier medical school in India[11] and 1.12 times more in Bench-Sheko Zone, Southwest Ethiopia[2].

These findings are almost 1.13 times lower than the study conducted in Indonesia[25], 1.12 times fewer than in Addis Ababa, Ethiopia[4],1.14 times less in Lagos, Nigeria[26], 1.15 times less in Kilimanjaro Region, Tanzania[27] and 1.2 times less in Rural Areas of Nigeria Sembilan, Malaysia[28] but consistent with the previous study conducted at the University of KwaZulu-Natal, South Africa[9].

The current national drive in Ethiopia for HPV vaccination may have contributed to the highest vaccine uptake rate in the study. During the data collection for this study, the parents were informed of the campaign-based school-based immunization program, and they decided on the vaccine. The high acceptability of the HPV vaccine in our study could be viewed as a chance to expand the country’s school-based HPV vaccination program. The study’s different acceptance rates in Addis Ababa, Gondar, and the Bench-Sheko Zone could be partially attributed to the respondents’ varied sociodemographic makeup.

Given that the nation has started a school-based vaccination program for these targeted girls, this shows that Ethiopia’s high parental desire to vaccinate their daughter is good news and presents an excellent opportunity as well. Because parents have a significant role in their daughters’ decision to receive the vaccine or not, a greater parental acceptance to have their daughter receives the HPV vaccine will facilitate the vaccination campaign by making more girls available for vaccination.

In this study, parents of daughters of the male sex were 59.3% less likely to accept HPV vaccination for their daughters as compared to the female sex (AOR: 0.407; 95%CI: 0.221, 0.748), which was almost similar to the study conducted a premier medical school in India[11], in Hong Kong[1], in Rural Areas of Nigeria Sembilan, Malaysia[28] and Bench-Sheko Zone, Southwest Ethiopia[2]. This could be because women are more likely to know about vaccine availability, the target group for immunization, and the catch-up campaign, and women may also be more willing to accept the vaccine and suggest it to others.

Those parents of daughter having a number of daughters one was 2.12 times more likely to accept HPV vaccination for their daughters as compared to the number of daughters with more than one (AOR: 2.122; 95%CI: 1.221, 3.685), this study found 1.57 times more than the study conducted in Shenzhen, china[12]. This could be in sum; many parents are willing but have not vaccinated daughters due to logistical barriers.

Parents of daughters having government school type were 52.4% less likely to accept HPV vaccination for their daughters as compared to private school (AOR: 0.476;95%CI: 0.263,0.861), this could be because private schools are funded wholly or partly vaccination of daughters for human papillomavirus vaccine by a private body.

Parents of daughters having poor knowledge were 46.8% less likely to accept HPV vaccination for their daughters as compared to good knowledge (AOR: 0.532; 95%CI: 0.293, 0.969), This study finding was consistent with the study conducted in Gondar town,

Northwest Ethiopia[18], almost 1.5 times more than the studies conducted in Bench-Sheko Zone, Southwest Ethiopia[2] and this study finding in contrast with the study finding in Arab communities[17].

This study finding is similar to the study conducted in Addis Ababa, Ethiopia[4], at a Teaching Hospital, Kuala Lumpur[29], in Kilimanjaro Region, Tanzania[27], but this study finding is also 2.82 times more than the study conducted in Lagos, Nigeria[24].

These disparities may have been brought about by the gap in their sociodemographic characteristics. This, in our opinion, demonstrates how parents’ acceptance to vaccinate their daughters is influenced by their knowledge of the HPV vaccination. Therefore, raising parental knowledge is crucial, and programs should be developed specifically for parents without a formal education.

Parents of daughters having negative attitude were 46% times less likely to accept HPV vaccination for their daughters as compared to positive attitudes (AOR: 0.540;95%CI: 0.299,0.977), Parental positive attitude towards HPV vaccine was significantly associated with parental acceptance to vaccinate their daughter. This is supported by other study findings[2], in Gondar town, Northwest Ethiopia[18]. This shows that more parents had a positive attitude toward the HPV vaccine despite their lack of understanding, and if vaccination services are offered, they would be willing to vaccinate their daughter.

## 5. Conclusion

This study found that there was a high level of parental acceptance to immunize their daughter against the human papillomavirus in the study area. Parents of daughters of the male sex, having multiple daughters, attending government schools, having poor knowledge, and having a negative attitude were discovered to have a substantial relationship with acceptance of the human papillomavirus vaccine. Additionally, the parents’ attitudes and knowledge about the HPV vaccine are significant in determining their intentions to vaccinate their daughters.

It is advised that there is a need to provide national educational campaigns about the HPV vaccine to the public given that the HPV vaccine is not included in the national immunization schedule. Authorities in areas where cervical cancer incidence is at risk should plan and implement by providing health information regarding the human papillomavirus vaccination with an emphasis on raising community awareness.

The regional and zonal health offices in Ethiopia’s ministry of health should develop health information regarding HPV vaccination with a focus on raising community awareness, especially among parents who lack formal education. It is essential to raise community awareness sustainably to secure the long-term acceptability of HPV vaccination. Therefore, it is essential to improve awareness and acceptance of cervical cancer as well as its prevention in the community.

## Limitation

To demonstrate the current parental acceptance to vaccinate their daughter in response to the vaccine campaign being run by the Ethiopian MOH and Zonal Health Department, a community-based study with a representative sample of parents from Ethiopia’s rural areas may be feasible. Additionally, it is the first study from a zonal level in Ethiopia to fully characterize parental understanding and acceptance of the HPV vaccination. Since the study is cross-sectional, a causal relationship cannot be shown; nonetheless, recollection and social desirability bias may potentially have an impact.

## Data Availability

All the minimal data set is made fully available without restriction

none

## Acknowledgments

Special thanks are extended to zonal, local educational department, local health managers, data collectors, supervisors, and Wachemo University.

## Consent

The author has consented to have their names and organizations acknowledged. Additionally, the material was collected and guaranteed anonymously to be confidential.

## Copyright

All authors must sign the “submission form” agreement before the article can be processed, and we all agreed to submit the manuscript to your journal.

## Notes

### Competing Interest Statement

The authors have declared no competing interest.

### Clinical Trial

none

### Clinical Protocols

none

### Funding Statement

The author(s) received no specific funding for this work

